# Acetaminophen and Acetaminophen-Opioid Combination Prescribing Trends Among Hospitalized Children, Adolescents, and Young Adults with Cancer

**DOI:** 10.1101/2025.05.08.25327260

**Authors:** Cheryl L. Mathis, Huong D. Meeks, Kevin M. Watt, Luke D. Maese, Jonathan E. Constance

## Abstract

**Purpose:** Acetaminophen (**APAP**) is a ubiquitous antipyretic and analgesic used in children in the United States (**US**), including those with cancer. The effects of US Food & Drug Administration (**FDA**) guidance on APAP prescribing have been described for healthy adults and children; however, APAP use patterns in children with cancer are unknown. Considering their increased risk of liver injury, APAP’s potential for causing hepatoxicity, and FDA guidance changes, this study examined the recent evolution of APAP use in children with cancer.

**Methods:** This retrospective, multi-center analysis extracted APAP prescribing data from the Pediatric Health Information System® (**PHIS**). Eligible children were aged 0-26 years, had a cancer diagnosis per International Classification of Diseases (**ICD**) codes, and were prescribed a chemotherapeutic. APAP and APAP-opioid combination prescribing were assessed at hospital, regional, and national levels. Changes in APAP and APAP-opioid combination use rates were assessed using the non-parametric Mann-Kendall test.

**Findings:** PHIS records for the complete years of 2004-2021 yielded 388,364 inpatient encounters for 50,779 unique patients. Of these, 87.3% of patients received APAP. Although APAP-opioid combination use was infrequent overall, children receiving APAP were more likely to receive an APAP-opioid combination medication (N=25,880, 13.4%, p < 0.001) compared to those who did not receive APAP. Among specialty children’s hospitals, national APAP use was stable over the study period. Regionally, APAP use increased among hospitals in the Northeast. APAP-opioid combination use decreased nationally with regional variation. In contrast to the steady decline in other regions, Southern APAP-opioid combination use was consistently elevated before declining in 2014.

**Implications:** This article describes acetaminophen and acetaminophen-opioid prescribing trends among children with cancer in the United States. These trends are key to help clinicians assess changes in pain management strategies over time, contextualize analgesic exposure and efficacy, and provide a foundation for future studies in drug safety. Extensive acetaminophen use can affect liver health, and further work is needed to evaluate acetaminophen exposure in children with cancer.

**Data Statement:** Deidentified data were obtained and evaluated under an IRB-approved protocol. Due to privacy requirements, the data are not available to be shared.

## Introduction

Analgesics are essential for managing pain and improving quality of life for children with cancer.^1^ In this population, pain is a frequently reported symptom arising from both malignancy and cancer therapy.^2–4^ In a 2020 study of hospitalized children with hematologic malignancies, the analgesic and antipyretic drug acetaminophen (**APAP**) was commonly used.^5^ However, prescribing rates of APAP and APAP-opioid combinations in a broad group of children with cancer in the United States (**US**) are poorly described. This knowledge gap is especially salient given the changing landscape of APAP safety and clinical guidance over the last 20 years.^6–8^ To evaluate the impact of safety guidance and gauge population-specific health risks, greater in-depth knowledge of APAP prescribing practices is needed.

Clinical use of APAP and APAP-opioid combinations is influenced by factors such as opioid prescribing regulations, pain severity, and the potential for toxicity.^9–13^ Indeed, APAP is a well-established hepatotoxin capable of inducing liver injury.^14–17^ Liver injury research has prompted the Food & Drug Administration (**FDA**) to issue multiple documents clarifying APAP’s risks and providing guidance for safe use (**Table 1**).^18–24^ Children with cancer are at an increased risk of acute liver injury primarily due to chemotherapy, radiation, or related sequelae (*e.g*., infection).^25–28^ In particular, chemotherapy often occupies the majority of clinical attention when considering hepatoxicity, with other supportive medications less frequently studied as contributors. Thus, it is vital to recognize APAP’s added hepatotoxicity poses a potential threat to children with cancer by inducing or exacerbating toxic liver outcomes.

**Table 1.**
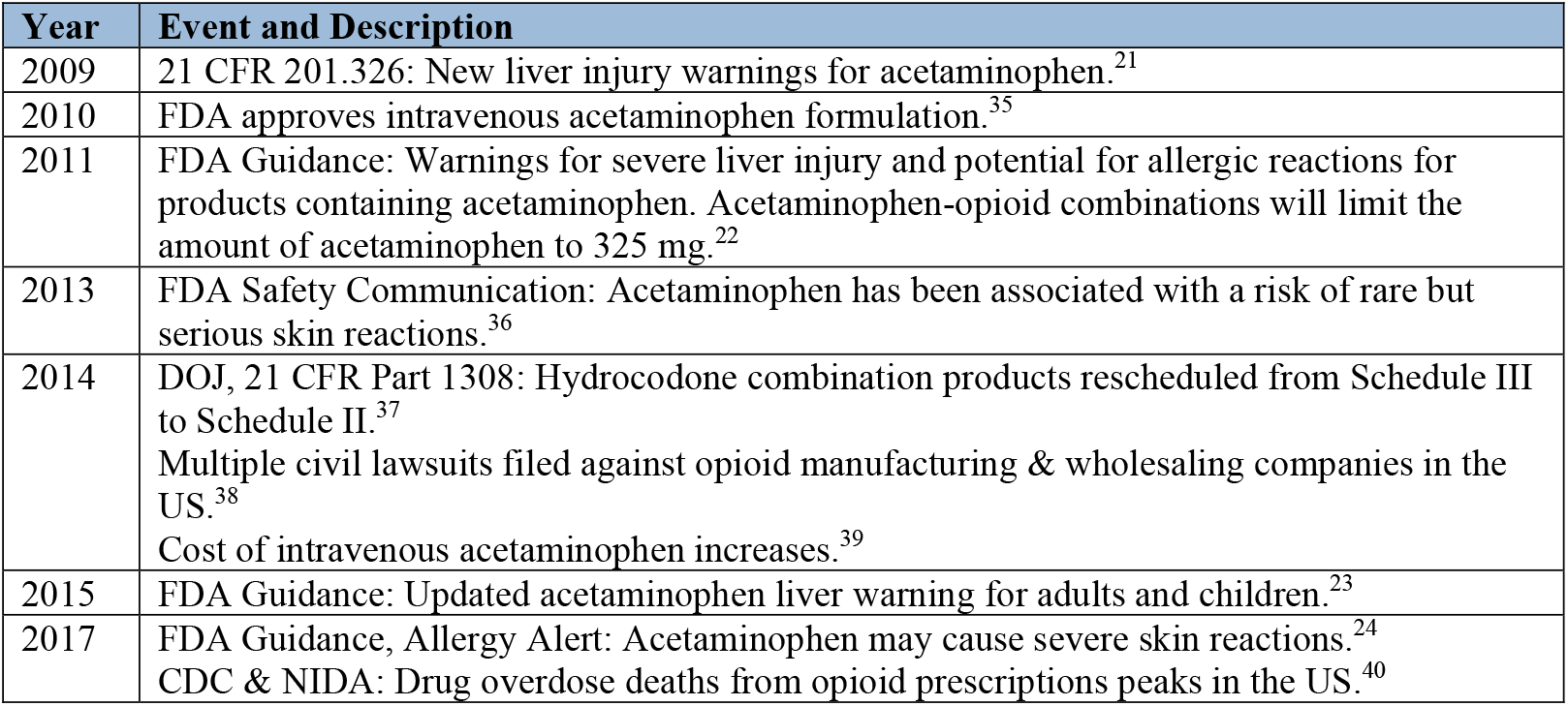
Regulatory and Social Events Surrounding APAP prescribing from 2004-2021.

| Year | Event and Description |
| --- | --- |
| 2009 | 21 CFR 201.326: New liver injury warnings for acetaminophen.<sup>21</sup> |
| 2010 | FDA approves intravenous acetaminophen formulation.<sup>35</sup> |
| 2011 | FDA Guidance: Warnings for severe liver injury and potential for allergic reactions for products containing acetaminophen. Acetaminophen-opioid combinations will limit the amount of acetaminophen to 325 mg.<sup>22</sup> |
| 2013 | FDA Safety Communication: Acetaminophen has been associated with a risk of rare but serious skin reactions.<sup>36</sup> |
| 2014 | DOJ, 21 CFR Part 1308: Hydrocodone combination products rescheduled from Schedule III to Schedule II.<sup>37</sup><br>Multiple civil lawsuits filed against opioid manufacturing & wholesaling companies in the US.<sup>38</sup><br>Cost of intravenous acetaminophen increases.<sup>39</sup> |
| 2015 | FDA Guidance: Updated acetaminophen liver warning for adults and children.<sup>23</sup> |
| 2017 | FDA Guidance, Allergy Alert: Acetaminophen may cause severe skin reactions.<sup>24</sup><br>CDC & NIDA: Drug overdose deaths from opioid prescriptions peaks in the US.<sup>40</sup> |

Additionally, when greater pain control is needed to manage cancer pain, APAP alone may prove ineffective.^29–31^ In these circumstances, APAP-opioid combinations may be prescribed.^32, 33^ Clinical APAP-opioid combination use is affected by opioid prescribing regulations, such as the Department of Justice’s (**DOJ**) 2014 reclassification of APAP-hydrocodone medications from Schedule III to Schedule II (**Table 1**).^8^ Further, the FDA implemented a 2014 mandate that APAP-opioid combinations may only contain up to 325 mg of APAP per dose to help reduce liver injury risk.^6^ Yet our understanding of how these regulatory changes have influenced and shifted clinical APAP use in children with cancer remains limited.

The majority of reports and reviews examining APAP and APAP-combination usage in children with cancer focus on immediate efficacy outcomes.^30, 32, 33^ Considering the heightened risk of liver injury in children with cancer, data that link APAP exposure, dosing, and prescribing are needed to understand its potential role in acute and late liver events. Therefore, this study sought to quantitatively describe APAP prescribing trends nationally and regionally using records in the Pediatric Health Information System® (**PHIS**). The PHIS database enabled further analysis at both the encounter and hospital levels. With changes in regulatory guidance from the FDA and DOJ, as well as changes in prescribing practices in response to the opioid crisis, we hypothesized that APAP use would remain prevalent while APAP-opioid combination use would decline.^6–8, 34^

## Participants and Methods

### PHIS Hospital and Patient Cohort

The PHIS database, supported by the Children’s Hospital Association, collects both clinical and resource utilization data from children’s hospitals across the US.^41^ This study was a retrospective, multicenter analysis of APAP prescribing trends using deidentified PHIS data conducted under an Intermountain Health and University of Utah Institutional Review Board (**IRB**)-approved protocol with waived consent. This study adhered to reporting guidelines from Consolidated Standards of Reporting Trials (**CONSORT**) and Strengthening the Reporting of Observational Studies in Epidemiology (**STROBE**).^42,43^ Owing to the retrospective nature of this analysis, there was no patient or public involvement in this study’s design or conduct.

Hospital encounter data, including medications administered during the encounter, were extracted from PHIS for the complete years of 2004-2021. Included patients were aged 0-26 years, had a cancer diagnoses code, were prescribed at least one of 95 pre-identified chemotherapeutics drugs (**SI Table 1**), and were treated at a PHIS hospital that had a 100% contribution rate for all quarters in the study period. Reporting to PHIS by contributing institutions occurs on a quarterly basis. Cancer diagnoses were identified based on International Classification of Diseases (**ICD**) codes (ICD-9 140-239 or ICD-10 C0-D4).^5^ The following exclusion criteria were applied: 1) non-inpatient encounters, and 2) encounters with length of stay of less than two days. For the purposes of this study, “encounter” was defined to be a period of inpatient clinical care. Unique patients were designated using a combination of medical record number (**MRN**), date of birth, and sex assignment provided in PHIS.

### APAP Use

APAP use was identified using generic drug titles available in PHIS. Single-component APAP was defined as drugs listed under “Acetaminophen (APAP) (*N*-Acetyl-*p*-aminophenol),” while APAP-opioid combinations were listed under “Acetaminophen combinations.” Throughout this study, APAP refers to single-component APAP unless otherwise specified. Patients receiving APAP as a single component drug and/or APAP formulated with an opioid (APAP-opioid combinations) *via* any route of administration were included. All routes of administration (*e.g*., intravenous, feeding tube) were included to ensure APAP use among children unable to tolerate oral dosing was captured.

### Statistical Analysis

Characteristics of both eligible patients and hospital encounters were summarized using counts (n, %) for categorical variables and medians (interquartile range, **IQR**) for continuous variables. Using Pearson’s Chi-squared tests and Fisher’s exact tests for categorical variables and Wilcoxon rank sum tests for continuous variables, data for both patient demographics and encounter-level characteristics were compared when APAP was/was not administered. Annual rates of APAP use and APAP-opioid combination use, defined as number of days administered per 1,000 hospital days, were calculated at the national, regional, and hospital-specific levels using **Equation 1**.

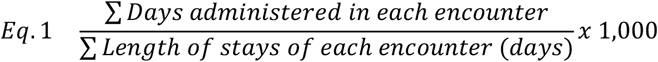

For our regional analysis, Centers for Disease Control (**CDC**) maps were used to group hospitals into four categories: Midwest, Northeast, South, and West.^44^ Encounters were assigned to a specific year based on the patient’s hospital admission date. The national-level data calculations included data from all PHIS hospitals eligible for analysis, while regional calculations included data from PHIS hospitals specific to each respective region. Changes in the rates of APAP use and APAP-opioid combination use over time were assessed at the hospital, regional, and national levels using the non-parametric Mann-Kendall trend test. Significance was defined as *p* < 0.05.

All analyses were completed using R, version 4.3.2 (R Project for Statistical Computing).^45, 46^

## Results

Out of 47 contributing PHIS hospitals, the 100% contribution rate requirement yielded 25 PHIS hospitals across 17 states. This included 10 hospitals in the Midwest, 2 in the Northeast, 8 in the South, and 5 in the West. A total of 388,364 inpatient encounters were eligible for analysis, representing 50,779 unique patients (**Scheme 1**).

Demographic results for the study population are shown in **Table 2**. The majority of children with cancer were designated as male (56.3%) and non-Hispanic white (54.6%) in PHIS; this study population reflected demographics seen in other US studies of children with cancer.^47, 48^ In the study group, single-component APAP was administered to 87.3% of children (N=44,348). Encounter characteristics are described **Table 3**. Except for the young adult group (18 to <27 years), APAP administration at a given encounter was at least as likely as receiving no APAP. The overall median length of stay was 4.0 days (IQR: 3.0, 8.0), while encounters involving APAP administration were longer (median, [IQR]: 6.0 days, [4.0, 13.0], *p* < 0.001). Encounters for patients 12 years of age and older were more frequent (Table 3). A sub-analysis of patient length of stay by age is included in **SI Table 2**: overall, length of stay decreased for patients 12 years and older. Encounters with APAP use were also more likely to include an APAP-opioid combination prescription (13.4%, *p* < 0.001) compared to those without APAP. Additionally, APAP use was associated (*p* < 0.001) with surgical operations (43.7%), surgical complications (16.2%), infections (33.9%), and total parenteral nutrition (**TPN**, 16.8%).

**Scheme 1.**
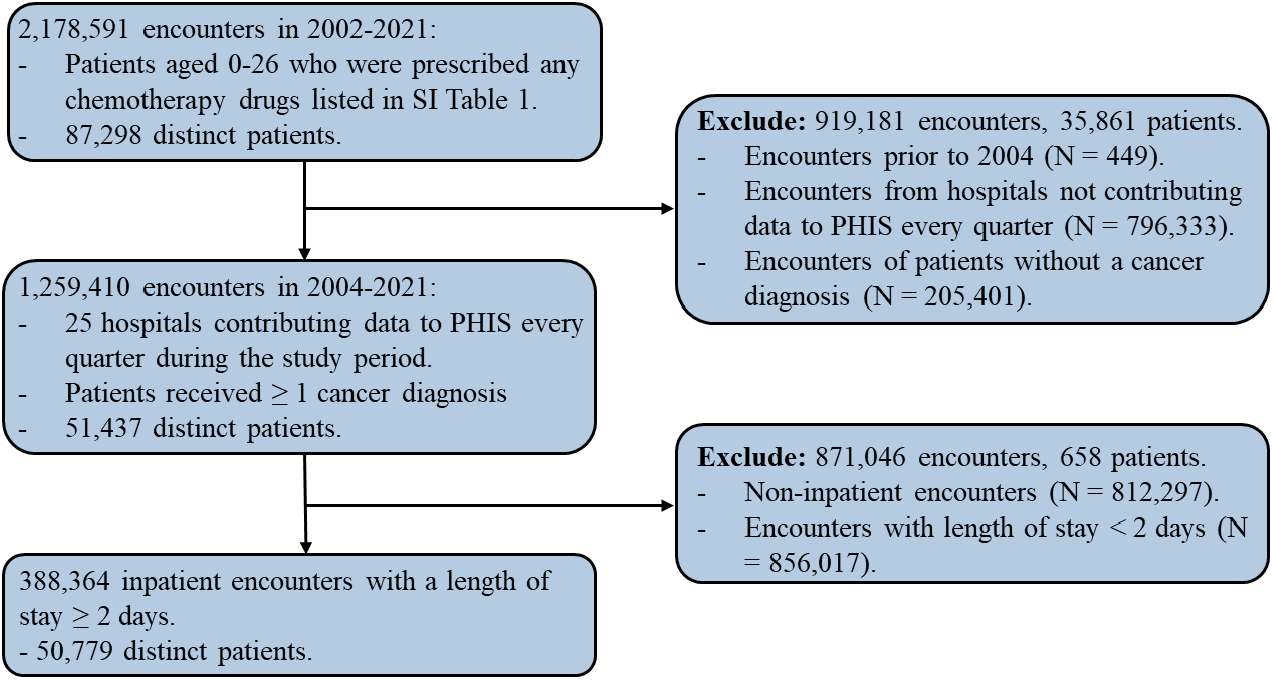
CONSORT Diagram for PHIS Analysis of APAP prescribing in children with cancer.

**Table 2.**
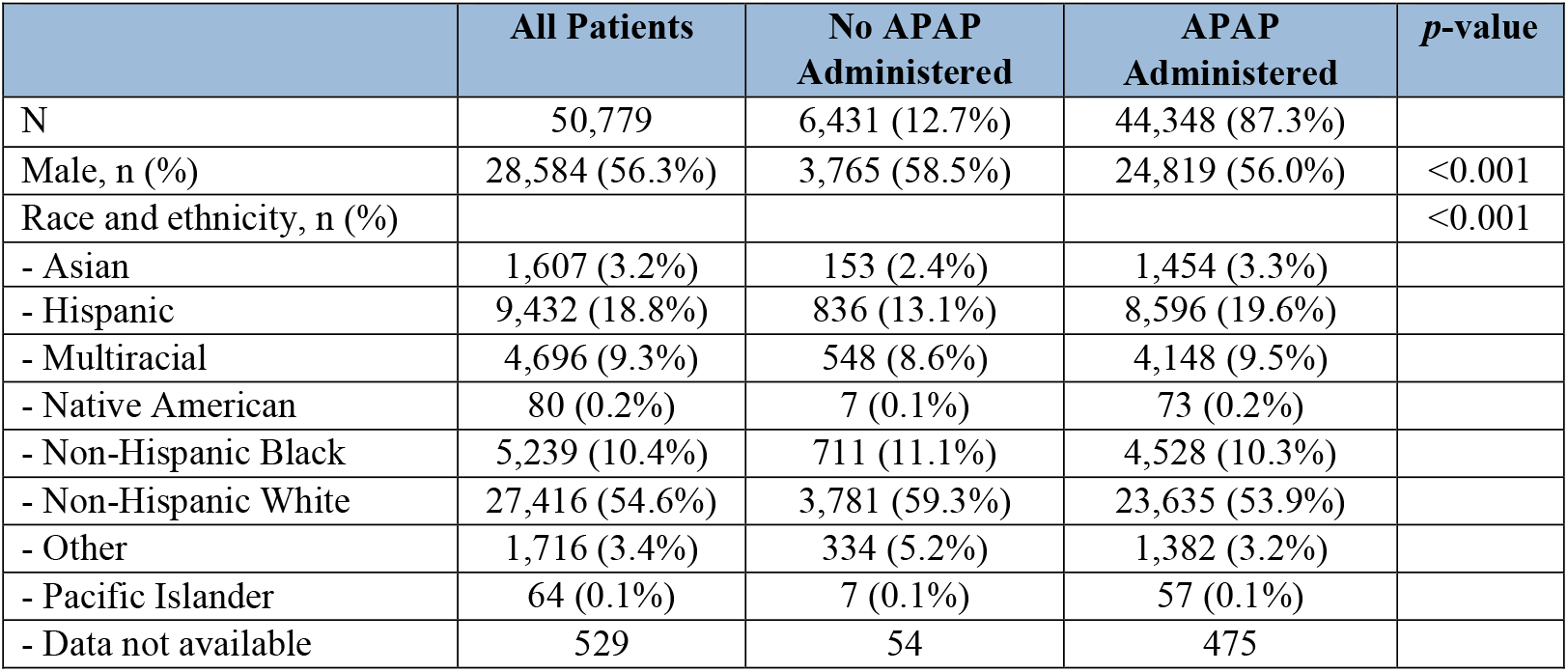
Patient-level demographics for children with cancer at PHIS hospitals from 2004-2021.

**Table 3.** Encounter-level characteristics of children with cancer at PHIS hospitals from 2004-2021.

|  | All Encounters | No APAP Administered | APAP Administered | *p*-value |
| --- | --- | --- | --- | --- |
| N | 388,364 | 195,890 (50.4%) | 192,474 (49.6%) |  |
| Age at admission |  |  |  | <0.001 |
| 0-27 days | 2,200 (0.4%) | 1,125 (0.4%) | 1,075 (0.4%) |  |
| 28 days- <2 years | 1,477 (0.4%) | 768 (0.4%) | 709 (0.4%) |  |
| 2- <12 years | 39,141 (10.1%) | 17,750 (9.1%) | 21,391 (11.1%) |  |
| 12- <18 years | 201,924 (52.0%) | 96,459 (49.2%) | 105,465 (54.8%) |  |
| 18- <27 years | 111,340 (28.7%) | 62,171 (31.7%) | 49,169 (25.5%) |  |
| Length of stay (days) | 4 (3, 8) | 4 (3, 5) | 6 (4, 13) | <0.001 |
| APAP-opioid combination administered | 38,397 (9.9%) | 12,517 (6.4%) | 25,880 (13.4%) | <0.001 |
| TPN charge | 44,367 (11.4%) | 11,971 (6.1%) | 32,396 (16.8%) | <0.001 |
| Operating room charge | 127,273 (32.8%) | 43,257 (22.1%) | 84,016 (43.7%) | <0.001 |
| Infection charge | 93,783 (24.1%) | 28,505 (14.6%) | 65,278 (33.9%) | <0.001 |
| Surgical complication | 45,962 (11.8%) | 14,754 (7.5%) | 31,208 (16.2%) | <0.001 |
| Renal condition | 44,054 (11.3%) | 15,689 (8.0%) | 28,365 (14.7%) | <0.001 |
| Transplant condition<sup>a</sup> | 28,067 (7.2%) | 8,895 (4.5%) | 19,172 (10.0%) | <0.001 |

<sup>a</sup>Feudtner's complex chronic condition.<sup>49</sup>

**Figure 1A** shows the annual rate variation of APAP use from 2004 to 2021 across hospitals, with a bolded black line depicting median rate. Across eligible PHIS hospitals, APAP use varied widely; however, the median fluctuated between 200 and 300 days of use per 1,000 hospital days. **Figure 1B** shows the annual rate of APAP use from 2004 to 2021 by region and nationally. Nationally, the trend in APAP use remained largely stable, with minor fluctuations over the study period (τ = 0.31, *p* = 0.081). The gray dashed vertical lines in **Figure 1B** indicate years when the FDA issued safety guidance for APAP use (Table 1). Both specialty children’s hospitals in the Northeast region showed an increase in APAP use, with APAP use rate more than doubling between 2004 and 2021 (rate = 84.5 days of APAP use per 1,000 hospital days in 2004 *vs*. 163.7 in 2021, τ = 0.61, *p* < 0.001). In the Midwest, APAP use decreased slightly (τ = −0.48, *p* = 0.005). There was no significant change in APAP use in the South (τ = 0.07, *p* = 0.709) or the West (τ = 0.32, *p* = 0.069).

**Figure 1.**
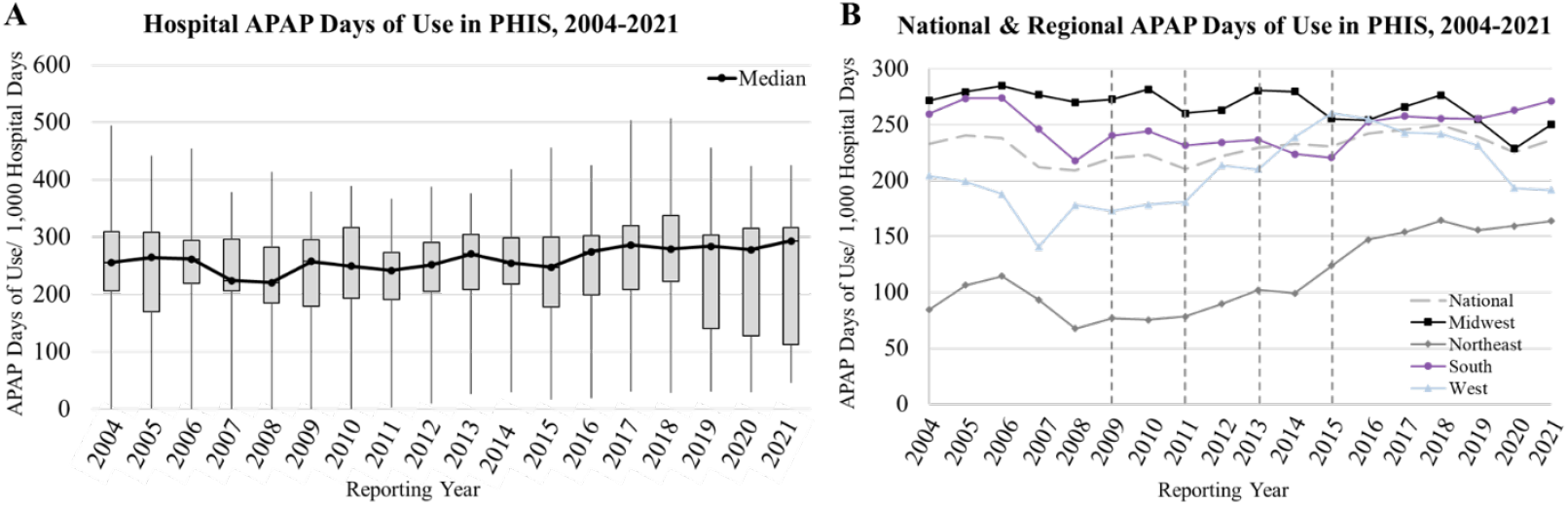
APAP Days of Use in PHIS. **(A)** Annual aggregate summary for all included PHIS hospitals. **(B)** National & regional median days of APAP use. Vertical gray lines in **B** correspond to regulatory events in 2009, 2011, 2013, and 2015 (Table 1).

The trend of APAP-opioid combination use shown in **Figure 2** contrasts with single-component APAP (Figure 1). While the national rate of APAP use remained stable, APAP-opioid combination use decreased over the study period (τ = −0.88, *p* < 0.001). In the Northeast, the observed decrease in APAP-opioid combination use coincided with an increase in APAP use (τ = − 0.91, *p* < 0.001). A similar trend was observed in the Midwest (τ = −0.97, *p* < 0.001). In the West, APAP-opioid use was stable until a decline began in 2016 (τ = −0.53, *p* < 0.002). Finally, the Southern APAP-opioid use rate peaked for the last time for the study period in 2014 at 74.8 days of use per 1,000 hospital days prior to decreasing (τ = −0.33, *p* = 0.057).

**Figure 2.**
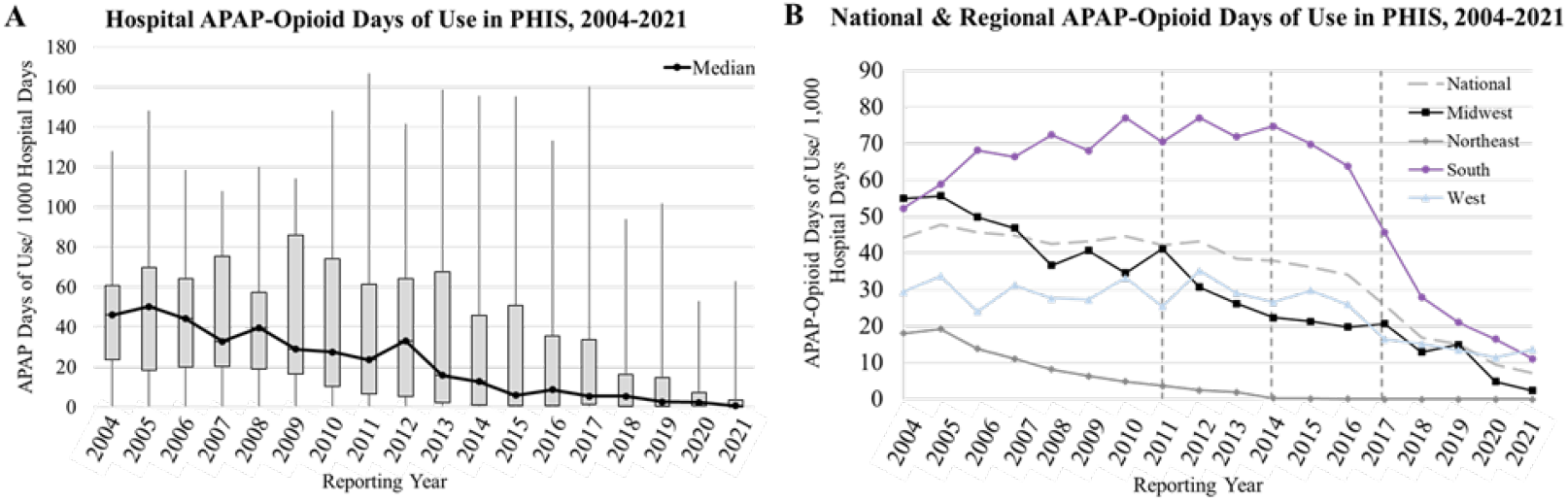
APAP-Opioid Combination Days of Use in PHIS. **(A)** Annual aggregate summary for all included PHIS hospitals. **(B)** National & regional median days of APAP-opioid combination use. Vertical gray lines in **B** correspond to regulatory events listed in 2011, 2014, and 2017 (Table 1).

Although the single-component APAP use rate remained stable when analyzed at the national level, 16 of the 25 hospitals had fluctuating use over time. Of these hospitals, 11 demonstrated significant increases in APAP use, while 5 showed significant decreases (See **SI Table 3**). The hospital-level rates of APAP use are shown graphically in **SI Figures 2A-C** for hospitals with increasing, mixed, and decreasing APAP use patterns. In contrast, all 18 hospitals demonstrated significant decreases in APAP-opioid combination use rates in 2021 compared to 2004. The hospital-level rates of APAP-opioid combination use are shown graphically in **Figures SI 3A-B** for hospitals with mixed and decreasing use patterns.

## Discussion and Conclusions

### National Analysis

Using the PHIS database, this study analyzed APAP prescribing trends for children with cancer in the US from 2004-2021. Nationally, APAP prescribing trends in this population remained stable, with days of APAP use per 1,000 hospital days ranging from 232.7 in 2004 to 236.1 in 2021. The median hospital length of stay was longer for children who received APAP (6 days) as compared to those who did not (4 days). Age at admission encounter characteristics in Table 3 demonstrated a high admission rate for children aged 12 years and older. An examination of hospital length of stay by age demonstrated that average length of stay largely decreased with increasing age: shorter but more frequent inpatient visits may account for the high rate of admission seen for older children.

Among all children with cancer diagnoses, 87.3% were prescribed APAP during at least one inpatient encounter. A previous study by Biltaji *et al*. reported APAP use for a single institution at 71.3% among children with hematologic malignancies who were also administered an antimicrobial.^5^ However, differences in study design limit comparison with our observed national and regional trends. The high rate of APAP use found in this study is complemented by the low use of non-steroidal anti-inflammatory drugs (**NSAIDs**) in children with cancer reported by Shen *et al*.^50^ In their report, only 9% of children with cancer received NSAIDs. NSAIDs pose significant risks for children with hematologic malignancies and other cancers due to their antiplatelet effects. Similarly, APAP is both an antipyretic and analgesic, thus its use must be carefully considered in children with cancer to avoid masking fever and underlying infections.

APAP’s dual functionality may partially account for its extensive use: antipyretic indications are less likely to be affected by changes in FDA guidance for liver injury. Both APAP and NSAIDs require thoughtful consideration prior to use; however, the stability and prevalence of APAP use found herein reflect consistent use as a cornerstone analgesic for children with cancer.

This study also found that children who received APAP were more likely to receive an APAP-opioid combination (APAP formulated together with an opioid drug). Importantly, APAP-opioid combination use among children with cancer was less prevalent as compared to APAP; indeed, over the study period the rate of APAP-opioid combination use declined. Nationally, APAP-opioid combination use substantially decreased from a median of 45 days per 1,000 hospital days in 2004 to less than 1 day in 2021. Based on an analysis of opioid prescribing from Shen *et al*., 71% of children with cancer received an opioid. Taken together, these findings may suggest a clinical shift away from APAP-opioid combination drugs toward single-component opioid drugs. In addition to efforts to curtail opioid misuse, this trend may be influenced by drug label changes, regulatory warnings, and reformulation strategies introduced by the FDA to decrease liver failure outcomes associated with APAP use.^21–24^

### Regional Analysis

Across three of four US regions, APAP use was stable (Midwest, South, and West). In the Northeastern US, the APAP prescribing rate increased (2004: 84.5/1,000 hospital days; 2021: 163.7/1,000 hospital days). APAP-opioid combination use exhibited varying trends but culminated in an overall decrease in prescribing. Both the Midwest and Northeast demonstrated a consistent decline throughout the study period, whereas trends in the West and South demonstrated small fluctuations until about 2015-2016. Western hospitals’ APAP-opioid combination use was found to be largely stable until 2016, after which it decreased. Southern US hospitals’ APAP-opioid combination use peaked at 77.1 days of use per 1,000 hospital days in both 2010 and 2012, before starting its decline in 2014 (Figure 2B). The 2014 decline may reflect multiple social and regulatory changes that year (Table 1), including the rescheduling of hydrocodone, the 325 mg cap of APAP in opioid combination products, and the multiple lawsuits filed against opioid manufacturers and wholesalers. The South’s sharp decline in APAP-opioid combination prescribing may also reflect concerns over rising synthetic opioid-related fatalities in Alabama, Florida, Louisiana, and North Carolina between 2015 and 2016, as reported by Cooper *et al*.^51^ Unfortunately, data from all five states most severely impacted by the opioid crisis (West Virgina, New Hampshire, Ohio, Maryland, and Massachusetts) were not available to further test this rationale.^51^

Limitations in this analysis include potential discrepancies between encounters reported in PHIS and actual encounters. This study employed a 100% PHIS contribution threshold (*i.e*., all included hospitals report data to PHIS every quarter) which restricted the number of hospitals eligible for analysis. While the 100% threshold data provide high-quality insights into APAP prescribing patterns, the smaller sample size in some regions (*e.g*., the Northeast) may not fully reflect APAP use trends at specialty children’s hospitals. Finally, specific APAP dose quantities and complete laboratory results were not available using PHIS data.

This study also demonstrates relationships between APAP use, duration of hospital stays, and key events (*e.g*., surgery and infection). However, these relationships cannot be unequivocally tied to clinical hepatotoxicity risk or outcomes in children with cancer using PHIS data. It is possible that even in the setting of mild to moderate liver toxicity, APAP may be used in children with cancer. Further quantitative studies of liver biomarkers (*e.g*., liver function tests and APAP-protein adducts) are needed to deconvolute liver injury from APAP-induced liver injury.^17, 52–55^ More granular data are needed to associate elevated liver enzymes and APAP dosing across encounters to describe APAP’s potential role in liver injury.

Overall, this study provides quantitative national and regional insights into APAP prescribing trends for children with cancer in the US. Using a retrospective, multicenter analysis of PHIS data, we observed a robust and steady rate of national APAP use in children with cancer.

Our findings suggest that while APAP use remained stable nationally, the use of comparatively rare APAP-opioid combinations declined. Although this analysis was specific to children with cancer, all cancer diagnoses were included to enhance population generalizability. Given the extensive use of APAP, further studies are needed to examine how APAP prescribing relates to potential toxicity (*e.g*., dosing and duration of use). To assess APAP’s contribution to liver sequalae in children with cancer, significant efforts are still needed to link prescribing data with quantitative drug exposure and liver function metrics.

## Data Availability

Deidentified data were obtained and evaluated under an IRB-approved protocol. Due to privacy requirements, the data are not available to be shared.

## Acknowledgments

The authors would like to thank Alfredo Novoa and Kelly Huynh of Pediatric

Enterprise Analytics, Intermountain Children’s Health. The authors thank the National Institutes of Health for project funding: NIH1K22CA258671-01A1 (JEC) and NIH NCATS CCTS-PCHF-002580 (JEC).

