## Supplementary Information for "Acetaminophen and Acetaminophen-Opioid Combination Prescribing Trends Among Hospitalized Children, Adolescents, and Young Adults with Cancer"

### Index

**SI Table 1:** List of 95 Chemotherapeutic Drugs Used to Screen PHIS Data. SI 3

**SI Table 2:** Hospital Encounter Length of Stay by Age Sub-Analysis SI 3

**SI Table 3:** Test for Monotonic Trends of APAP Use Using Mann-Kendall Test. SI 4

**SI Figure 1:** APAP Use Among Hospitals Identified Using Kendall's Correlation. SI 5

**SI Figure 2:** APAP-Opioid Combination Use Among Hospitals Identified Using Kendall's Correlation. SI 5

**SI Table 4:** CONSORT Checklist. SI 6

**SI Tabl5 5:** STROBE Checklist. SI 10

**SI Table 1:** List of 95 Chemotherapeutic Drugs Used to Screen PHIS Data

| | | | | |
|---|---|---|---|---|
| Ifosfamide | Docetaxel | Trastuzumab | Carboplatin | Dasatinib |
| Trabectedin | Eribulin | Pertuzumab | Cisplatin | Erlotinib |
| Lomustine | Daunorubicin | Obinutuzumab | Oxaliplatin | Gefitinib |
| Streptozocin | Doxorubicin | Blinatumomab | Procarbazine | Sorafenib |
| Dacarbazine | Epirubicin | Dinutuximab | Ponatinib | Vemurafenib |
| Bendamustine | Bleomycin | Siltuximab | Palbociclib | Arsenic trioxide |
| Pralatrexate | Idarubicin | Ibritumomab tiuxetan | Temozolomide | Axitinib |
| Capecitabine | Dactinomycin | Tositumomab | Azacitidine | Vismodegib |
| Clofarabine | Mitomycin | Pembrolizumab | Irinotecan | Carfilzomib |
| Cytarabine | Romidepsin | Daratumumab | Topotecan | Regorafenib |
| Gemcitabine | Bicalutamide | Elotuzumab | Aldesleukin (interleukin-2) | Omacetaxine mepesuccinate |
| Nelarabine | Flutamide | Olaratumab | Crizotinib | Trametinib |
| Fludarabine phosphate | Fulvestrant | Inotuzumab ozogamicin | Denileukin diftitox | Dabrafenib |
| Pemetrexed | Anastrozole | Etoposide | Pazopanib | Pegaspargase (PEG-L-asparaginase) |
| Thioguanine | Exemestane | Ceritinib | Nilotinib | |
| Vinblastine | Gemtuzumab ozogamicin | Teniposide (VM-26) | Bexarotene | |
| Vincristine | Panitumumab | Panobinostat | Lapatinib | |
| Venetoclax | Cetuximab | Vorinostat | Sunitinib | |
| Vinorelbine | Ofatumumab | Asparaginase | Temsirolimus | |
| Paclitaxel | Ipilimumab | Ribociclib | Imatinib | |

**SI Table 2:** Hospital Encounter Length of Stay by Age Sub-Analysis.

| Age at admission | N | Min | Max | Median | Q1 | Q3 | Mean | SD |
|---|---|---|---|---|---|---|---|---|
| - 0-27 days | 1477 | 2 | 746 | 16 | 5 | 42 | 37.4 | 60.0 |
| - 28 days-<2 years | 39141 | 2 | 1103 | 5 | 3 | 11 | 11.5 | 22.1 |
| - 2- <12 years | 201924 | 2 | 1413 | 4 | 3 | 8 | 8.2 | 13.8 |
| - 12- <18 years | 111340 | 2 | 403 | 4 | 3 | 7 | 8.1 | 13.4 |
| - 18- <27 years | 34482 | 2 | 510 | 4 | 3 | 8 | 8.9 | 15.0 |

**SI Table 3:** Test for Monotonic Trends of APAP Use Using Mann-Kendall Test.

| Hospital ID | APAP | | APAP-Opioid Combination | |
|---|---|---|---|---|
| | rho | p-value | rho | p-value |
| MW1 | -0.41 | **0.017** | -0.75 | **0.000** |
| MW2 | 0.32 | **0.068** | -0.62 | **0.000** |
| MW3 | 0.40 | **0.021** | -0.91 | **0.000** |
| MW4 | -0.05 | 0.823 | -0.75 | **0.000** |
| MW5 | 0.10 | 0.601 | -0.58 | **0.000** |
| MW6 | 0.45 | **0.009** | -0.75 | **0.000** |
| MW7 | -0.10 | 0.601 | -0.83 | **0.000** |
| MW8 | -0.58 | **0.000** | -0.78 | **0.000** |
| MW9 | -0.65 | **0.000** | -0.79 | **0.000** |
| MW10 | 0.45 | **0.009** | -0.24 | 0.175 |
| NE1 | 0.80 | **0.000** | -0.97 | **0.000** |
| NE2 | 0.61 | **0.000** | -0.41 | **0.042** |
| S1 | 0.42 | **0.014** | -0.76 | **0.000** |
| S2 | 0.08 | 0.654 | -0.18 | 0.320 |
| S3 | 0.63 | **0.000** | -0.65 | **0.000** |
| S4 | -0.63 | **0.000** | 0.07 | 0.709 |
| S5 | 0.69 | **0.000** | -0.46 | **0.007** |
| S6 | -0.33 | **0.057** | -0.71 | **0.000** |
| S7 | -0.20 | 0.260 | -0.19 | 0.293 |
| S8 | 0.56 | **0.001** | -0.56 | **0.002** |
| W1 | 0.24 | 0.175 | -0.69 | **0.000** |
| W2 | 0.42 | **0.014** | 0.27 | 0.131 |
| W3 | 0.49 | **0.004** | -0.06 | 0.765 |
| W4 | -0.45 | **0.009** | -0.78 | **0.000** |
| W5 | 0.42 | **0.014** | -0.02 | 0.941 |

MW: Midwest; NE: Northeast; S: South; W: West. Bolded text highlights instances in which $p < 0.05$, the threshold of significance.

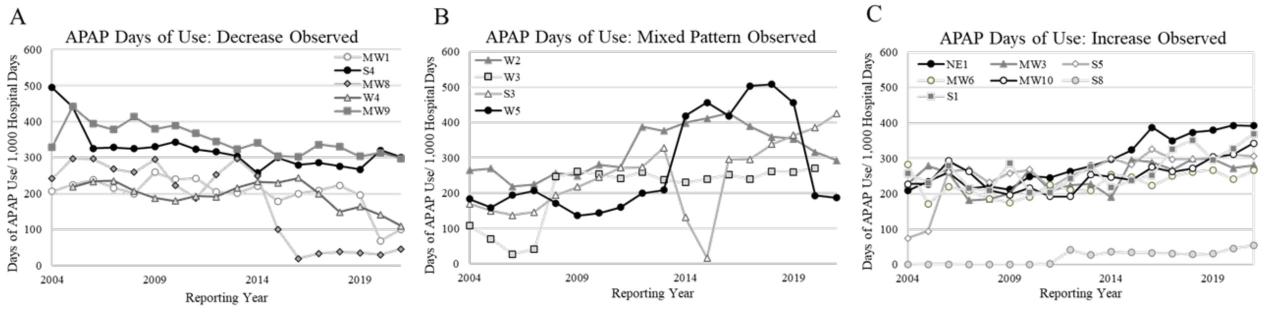


**SI Figure 2:** APAP Use Among Hospitals Identified Using Kendall's Correlation. **A.** Decreasing APAP use; **B.** Mixed Increasing-Decreasing APAP use; **C.** Increasing APAP use observed. MW: Midwest; NE: Northeast; S: South; W: West.

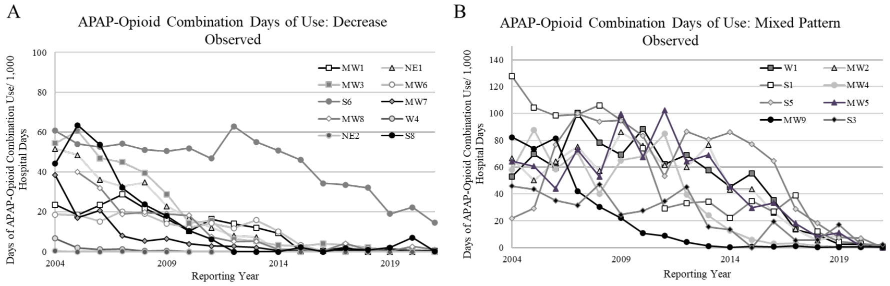


**SI Figure 3:** APAP-Opioid Combination Use Among Hospitals Identified Using Kendall's Correlation. **A.** Decreasing APAP use and **B.** Mixed Increasing-Decreasing APAP use observed. MW: Midwest; NE: Northeast; S: South; W: West.

**Table 4:** CONSORT Checklist.

### CONSORT 2010 checklist of information to include when reporting a randomised trial*

| Section/Topic | Item No | Checklist item | Reported on page No |
|---|---|---|---|
| **Title and abstract** | | | |
| | 1a | Identification as a randomised trial in the title | NA |
| | 1b | Structured summary of trial design, methods, results, and conclusions (for specific guidance see CONSORT for abstracts) | 3 |
| **Introduction** | | | |
| Background and objectives | 2a | Scientific background and explanation of rationale | 4 |
| | 2b | Specific objectives or hypotheses | 5 |
| **Methods** | | | |
| Trial design | 3a | Description of trial design (such as parallel, factorial) including allocation ratio | NA |
| | 3b | Important changes to methods after trial commencement (such as eligibility criteria), with reasons | NA |
| Participants | 4a | Eligibility criteria for participants | 6 |
| | 4b | Settings and locations where the data were collected | 6 |
| Interventions | 5 | The interventions for each group with sufficient details to allow replication, including how and when they were actually administered | NA |

| | | | |
|---|---|---|---|
| Outcomes | 6a | Completely defined pre-specified primary and secondary outcome measures, including how and when they were assessed | NA |
| | 6b | Any changes to trial outcomes after the trial commenced, with reasons | NA |
| Sample size | 7a | How sample size was determined | 9 |
| | 7b | When applicable, explanation of any interim analyses and stopping guidelines | NA |
| Randomisation: | | | |
| Sequence generation | 8a | Method used to generate the random allocation sequence | NA |
| | 8b | Type of randomisation; details of any restriction (such as blocking and block size) | NA |
| Allocation concealment mechanism | 9 | Mechanism used to implement the random allocation sequence (such as sequentially numbered containers), describing any steps taken to conceal the sequence until interventions were assigned | NA |
| Implementation | 10 | Who generated the random allocation sequence, who enrolled participants, and who assigned participants to interventions | NA |
| Blinding | 11a | If done, who was blinded after assignment to interventions (for example, participants, care providers, those assessing outcomes) and how | NA |
| | 11b | If relevant, description of the similarity of interventions | NA |
| Statistical methods | 12a | Statistical methods used to compare groups for primary and secondary outcomes | NA |
| | 12b | Methods for additional analyses, such as subgroup analyses and adjusted analyses | 7-8 |
| **Results** | | | |

| | | | |
|---|---|---|---|
| Participant flow (a diagram is strongly recommended) | 13a | For each group, the numbers of participants who were randomly assigned, received intended treatment, and were analysed for the primary outcome | NA |
| | 13b | For each group, losses and exclusions after randomisation, together with reasons | NA |
| Recruitment | 14a | Dates defining the periods of recruitment and follow-up | NA |
| | 14b | Why the trial ended or was stopped | NA |
| Baseline data | 15 | A table showing baseline demographic and clinical characteristics for each group | 9 |
| Numbers analysed | 16 | For each group, number of participants (denominator) included in each analysis and whether the analysis was by original assigned groups | 9-10 |
| Outcomes and estimation | 17a | For each primary and secondary outcome, results for each group, and the estimated effect size and its precision (such as 95% confidence interval) | 9-10 |
| | 17b | For binary outcomes, presentation of both absolute and relative effect sizes is recommended | NA |
| Ancillary analyses | 18 | Results of any other analyses performed, including subgroup analyses and adjusted analyses, distinguishing pre-specified from exploratory | 10-12 |
| Harms | 19 | All important harms or unintended effects in each group (for specific guidance see CONSORT for harms) | NA |
| **Discussion** | | | |
| Limitations | 20 | Trial limitations, addressing sources of potential bias, imprecision, and, if relevant, multiplicity of analyses | 14-15 |
| Generalisability | 21 | Generalisability (external validity, applicability) of the trial findings | 15 |
| Interpretation | 22 | Interpretation consistent with results, balancing benefits and harms, and considering other relevant evidence | 12-14 |

| | | | |
|---|---|---|---|
| **Other information** | | | |
| Registration | 23 | Registration number and name of trial registry | NA |
| Protocol | 24 | Where the full trial protocol can be accessed, if available | NA |
| Funding | 25 | Sources of funding and other support (such as supply of drugs), role of funders | 1 |

Citation: Schulz KF, Altman DG, Moher D, for the CONSORT Group. CONSORT 2010 Statement: updated guidelines for reporting parallel group randomised trials. BMC Medicine. 2010;8:18.

© 2010 Schulz et al. This is an Open Access article distributed under the terms of the Creative Commons Attribution License (http://creativecommons.org/licenses/by/2.0), which permits unrestricted use, distribution, and reproduction in any medium, provided the original work is properly cited.

*We strongly recommend reading this statement in conjunction with the CONSORT 2010 Explanation and Elaboration for important clarifications on all the items. If relevant, we also recommend reading CONSORT extensions for cluster randomised trials, non-inferiority and equivalence trials, non-pharmacological treatments, herbal interventions, and pragmatic trials. Additional extensions are forthcoming: for those and for up-to-date references relevant to this checklist, see www.consort-statement.org.

**Table 5:** STROBE Checklist.

| | Item No | Recommendation | Page No |
|---|---|---|---|
| **Title and abstract** | 1 | (*a*) Indicate the study's design with a commonly used term in the title or the abstract | 1 |
| | | (*b*) Provide in the abstract an informative and balanced summary of what was done and what was found | 3 |
| **Introduction** | | | |
| Background/rationale | 2 | Explain the scientific background and rationale for the investigation being reported | 4 |
| Objectives | 3 | State specific objectives, including any prespecified hypotheses | 5 |
| **Methods** | | | |
| Study design | 4 | Present key elements of study design early in the paper | 6 |
| Setting | 5 | Describe the setting, locations, and relevant dates, including periods of recruitment, exposure, follow-up, and data collection | 6 |
| Participants | 6 | (*a*) *Cohort study*—Give the eligibility criteria, and the sources and methods of selection of participants. Describe methods of follow-up<br><br>*Case-control study*—Give the eligibility criteria, and the sources and methods of case ascertainment and control selection. Give the rationale for the choice of cases and controls<br><br>*Cross-sectional study*—Give the eligibility criteria, and the sources and methods of selection of participants | 6 |
| | | (*b*) *Cohort study*—For matched studies, give matching criteria and number of exposed and unexposed<br><br>*Case-control study*—For matched studies, give matching criteria and the number of controls per case | NA |
| Variables | 7 | Clearly define all outcomes, exposures, predictors, potential confounders, and effect modifiers. Give diagnostic criteria, if applicable | 7 |
| Data sources/ measurement | 8* | For each variable of interest, give sources of data and details of methods of assessment (measurement). Describe comparability of assessment methods if there is more than one group | 7 |
| Bias | 9 | Describe any efforts to address potential sources of bias | 7 |

| | | | |
|---|---|---|---|
| Study size | 10 | Explain how the study size was arrived at | 9 |
| Quantitative variables | 11 | Explain how quantitative variables were handled in the analyses. If applicable, describe which groupings were chosen and why | 7 |
| Statistical methods | 12 | (*a*) Describe all statistical methods, including those used to control for confounding | 7 |
| | | (*b*) Describe any methods used to examine subgroups and interactions | 7 |
| | | (*c*) Explain how missing data were addressed | 6 |
| | | (*d*) *Cohort study*—If applicable, explain how loss to follow-up was addressed<br>*Case-control study*—If applicable, explain how matching of cases and controls was addressed<br>*Cross-sectional study*—If applicable, describe analytical methods taking account of sampling strategy | NA |
| | | (*e*) Describe any sensitivity analyses | |

**Results**

| | | | |
|---|---|---|---|
| Participants | 13* | (a) Report numbers of individuals at each stage of study—eg numbers potentially eligible, examined for eligibility, confirmed eligible, included in the study, completing follow-up, and analysed | 9 |
| | | (b) Give reasons for non-participation at each stage | 9 |
| | | (c) Consider use of a flow diagram | 9 |
| Descriptive data | 14* | (a) Give characteristics of study participants (eg demographic, clinical, social) and information on exposures and potential confounders | 9 |
| | | (b) Indicate number of participants with missing data for each variable of interest | 9 |
| | | (c) *Cohort study*—Summarise follow-up time (eg, average and total amount) | NA |
| Outcome data | 15* | *Cohort study*—Report numbers of outcome events or summary measures over time | 11,12 |
| | | *Case-control study*—Report numbers in each exposure category, or summary measures of exposure | NA |
| | | *Cross-sectional study*—Report numbers of outcome events or summary measures | NA |

| | | | |
|---|---|---|---|
| Main results | 16 | (*a*) Give unadjusted estimates and, if applicable, confounder-adjusted estimates and their precision (eg, 95% confidence interval). Make clear which confounders were adjusted for and why they were included | 9,10 |
| | | (*b*) Report category boundaries when continuous variables were categorized | 8 |
| | | (*c*) If relevant, consider translating estimates of relative risk into absolute risk for a meaningful time period | NA |
| Other analyses | 17 | Report other analyses done—eg analyses of subgroups and interactions, and sensitivity analyses | 8 |
| **Discussion** | | | |
| Key results | 18 | Summarise key results with reference to study objectives | 13 |
| Limitations | 19 | Discuss limitations of the study, taking into account sources of potential bias or imprecision. Discuss both direction and magnitude of any potential bias | 14 |
| Interpretation | 20 | Give a cautious overall interpretation of results considering objectives, limitations, multiplicity of analyses, results from similar studies, and other relevant evidence | 15 |
| Generalisability | 21 | Discuss the generalisability (external validity) of the study results | 15 |
| **Other information** | | | |
| Funding | 22 | Give the source of funding and the role of the funders for the present study and, if applicable, for the original study on which the present article is based | 1, 16 |

*Give information separately for cases and controls in case-control studies and, if applicable, for exposed and unexposed groups in cohort and cross-sectional studies.

**Note:** An Explanation and Elaboration article discusses each checklist item and gives methodological background and published examples of transparent reporting. The STROBE checklist is best used in conjunction with this article (freely available on the Web sites of PLoS Medicine at http://www.plosmedicine.org/, Annals of Internal Medicine at http://www.annals.org/, and Epidemiology at http://www.epidem.com/). Information on the STROBE Initiative is available at www.strobe-statement.org.
